# Association between coastal residence since birth and blood pressure among adolescents: a cross-sectional comparative study

**DOI:** 10.64898/2026.07.27.26359069

**Authors:** Irin Hossain, M M Aktaruzzaman, Md Abdullah Saeed Khan, Manzurul Haque Khan

## Abstract

**Background:** Salinity intrusion in coastal Bangladesh poses significant public health challenges, with millions exposed to elevated sodium through drinking water. While dietary salt intake is a well-established risk factor for hypertension, the impact of environmental salinity exposure on blood pressure in adolescents remains poorly understood. This study examined the association between coastal residence since birth and blood pressure among school-going adolescents in Bangladesh.

**Methods:** This cross-sectional comparative study was conducted between January and December 2014 among 1,056 adolescents residing in high-salinity (Dacope, Khulna) and low-salinity (Singair, Manikganj) areas since birth. Blood pressure measurements, anthropometric data, and information on sociodemographics, drinking water sources, dietary salt intake, family history of hypertension, and physical activity were collected. Linear regression analysis examined the effect of residence on blood pressure after adjusting for age, sex, family history of hypertension, extra salt intake, BMI, and physical activity level.

**Results:** Participants had an average age of 14.95±0.78 years, and 66.76% were male. Those living in high-salinity areas had significantly lower BMI (17.58±2.14 vs 22.56±1.46 kg/m^2^), higher underweight prevalence (23.67% vs 0.19%), and predominantly used pond water (60.98%) compared to deep tubewells (86.17%) in low-salinity areas. After adjustment for confounders, residence in high-salinity areas since birth was independently associated with an 8.74 mmHg increase in systolic blood pressure (95% CI: 7.22 to 10.26, p<0.001) but showed no significant association with diastolic blood pressure (β=-0.97 mmHg, 95% CI: -2.18 to 0.25, p=0.120). Sensitivity analysis using median quintile regression confirmed these findings.

**Conclusion:** Coastal residence since birth is independently associated with elevated systolic blood pressure in adolescents, highlighting a potential risk factor of hypertension in coastal populations.

## Introduction

Water-related crises, particularly salinity intrusion, pose significant environmental and public health challenges in Bangladesh [1]. As a deltaic country, Bangladesh spans 147,570 km^2^, with its coastal region covering approximately 29,000 km^2^ (20% of the country), where 53% of the area is affected by varying degrees of salinity [2–4]. People living in coastal regions rely heavily on rivers, ponds, and groundwater for drinking water [5], which is increasingly contaminated due to saltwater intrusion, tidal surges, and soil runoff [6]. Climate change has further exacerbated this issue, with rising sea levels and reduced freshwater inflow intensifying inland salinity [7]. This growing environmental crisis has raised concerns about its potential health impacts, particularly blood pressure and hypertension [8,9].

Salinity is primarily caused by dissolved salts, with sodium chloride (NaCl) accounting for 65.7% of total salinity[2,7]. The World Health Organization (WHO) recommends a maximum chloride concentration of 250 mg/L in drinking water[10], yet many coastal areas in Bangladesh exceed this limit [11]. Chronic exposure to high-salinity drinking water has been hypothesized to contribute to hypertension and cardiovascular diseases [12], particularly in vulnerable populations such as young adults [13]. While previous studies have explored the role of dietary salt in hypertension, emerging evidence suggests that environmental salt exposure, mainly through drinking water, may have additional health implications [1]. Mueller and colleagues projected ≥1 km of inland saltwater intrusion in 41 nations including Bangladesh by 2050 [6], making this an urgent public health issue.

Blood pressure is influenced by multiple factors, including genetics, diet, physical activity, and environmental exposures [14]. Hypertension in children and adolescents is a growing global concern, with prevalence rates between 1.66 to 14.53% worldwide [15]. Evidence suggests that childhood blood pressure levels track into adulthood, making early identification and intervention crucial [16]. High salt intake is a known risk factor for hypertension, as it leads to sodium retention, increased plasma volume, and arterial constriction [17]. While excessive dietary salt intake has been widely studied, research on the long-term effects of environmental salinity exposure on blood pressure, particularly in children, remains limited.

In Bangladesh, the coastal region experiences higher hypertension prevalence than inland areas, suggesting a possible association between environmental salinity and blood pressure levels [9]. However, research on this topic is still evolving, with some studies suggesting a direct link between high-salinity drinking water and increased blood pressure [17]. In contrast, others emphasize lifestyle factors such as diet and obesity[14]. Given the increasing salinity intrusion and its potential health consequences, further investigation is warranted to establish risk associations and develop public health interventions

This study aims to assess and compare blood pressure levels among school-going adolescents residing in high- and low-salinity areas of Bangladesh since birth. By evaluating the effect of long-term residence in coastal high salinity areas on blood pressure, this research seeks to provide critical insights into a potential inherent risk of hypertension and contribute to evidence-based strategies for hypertension prevention in coastal populations.

## Materials and Methods

### Study Design and Place

This cross-sectional comparative study assessed blood pressure among school-going adolescents residing in high- and low-salinity areas of Bangladesh between September and December 2014. The study was conducted in high schools at Bajua and Chunphuri villages of Dacope Upazila of Khulna District, a high-salinity area, and Joymontop village of Singair Upazila of Manikganj District, a low-salinity area. These locations were selected based on the salinity levels observed in the 2009 baseline survey data on national drinking water quality in Bangladesh [18].

### Study Population and Sampling

The study targeted school-going adolescents studying in classes VIII, IX, and X who were residing in their respective villages since birth. Sample size was determined using the online calculator Simple Interactive Statistical Analysis, SISA (SISA Web Calculator). The option for calculating the sample size to compare two sample means was used. Considering 95% Confidence Interval, 80% power, 5% error, and taking the average diastolic blood pressure of 56.2 ± 11.7 mmHg (±SD) and 53.7±10.2 mmHg from school children with high and low levels of sodium in drinking water, respectively [19]. The sample size was calculated to be 440. To accommodate possible non-responses, the sample size was inflated by 20% to yield the final number of 528. A total of 1056 respondents were interviewed; among them, 50% were from high-salinity areas, and the rest were from low-salinity areas. Adolescents who were willing to participate were included. Those with a history in-migration to the study villages and a diagnosis of hypertension were excluded. Exclusion of adults also removed older age as confounding factor in the potential association between coastal residence and increased blood pressure.

### Data collection instrument and method

Data were collected through face-to-face interviews with structured questionnaires. Participants were interviewed to collect information about socio-demographics, sources of drinking water, extra salt intake, intake of pickled fish, and family history of hypertension (father and/or mother, paternal grandfather, paternal grandmother, maternal grandfather, or maternal grandmother). Anthropometric and blood pressure measurements were taken from all participants.

#### Blood Pressure Measurement

Blood pressure was measured following standard protocols following standard guidelines [20] to ensure accuracy. Measurements were taken in a controlled environment after five minutes of rest, with participants seated and their right arm supported at heart level. Arm circumference was measured at the midpoint between the olecranon process and the acromion. Blood pressure readings were taken three times, and the average of these readings was used for analysis. Average systolic blood pressure was obtained by taking the average of three readings. A similar approach was followed for calculating average diastolic pressure.

#### Anthropometric Measurements

Anthropometric measurements included height, weight, and body mass index (BMI). Height and weight were recorded with participants wearing light clothing and no shoes to ensure accuracy. BMI was calculated using the formula BMI = weight (kg) / height (m^2^) and categorized into three groups based on the BMI for age chart of the World Health Organization (WHO) growth reference developed by De Onis et al [21].

#### Definition of Extra Salt Intake

Extra salt intake was assessed using two parameters: direct salt intake (additional salt added to food) and consumption of salt-preserved fish. If a participant reported consuming neither, they were classified as having no extra salt intake; otherwise, they were categorized as having extra salt intake. This classification helped evaluate the influence of dietary salt consumption on blood pressure differences between high- and low-salinity groups.

#### Category of physical activity

Physical activity was assessed using the International Physical Activity Questionnaire (IPAQ) questionnaire and was categorized into three groups: low, moderate, and high, using the IPAQ guidelines [22]. MET (Metabolic Equivalent of Task) was expressed as minutes per week. The values of walking, moderate-intensity, and vigorous-intensity activities were assigned MET levels of 3.3, 4.0, and 8.0, respectively. The total MET (minutes/week) was calculated as the cumulative sum of walking, moderate, and vigorous METs.

### Statistical Analysis

Data was analyzed using R Studio Version 2025.09.2. As there were no missing data, imputation was not required. Continuous data distribution was checked using histograms with normality curves. Descriptive statistics were presented as frequency (percentage), mean (Standard Deviation [sd]) and median (Interquartile Range[IQR]) as appropriate. The Pearson’s chi-square test was used for categorical variables, while the Welch two-sample t-test and the Wilcoxon rank sum test were used to compare continuous values by categorical variables with two categories. Pearson’s and Kanda’s tau correlation was run to detect the association between age, BMI, and the log of MET minutes. The effect of residence (i.e., high-salinity vs low-salinity areas) on blood pressure was examined by adjusting for age, sex, family history of blood pressure, extra salt intake, BMI, and physical activity level (measured as MET minutes) through linear regression analysis. A sensitivity analysis was also conducted using median quintile regression to check for the consistency of the effects of residence on blood pressure levels. Supplementary figures (S1 Fig and S2 Fig) show the results of the post-hoc assumptions checks for linear regression models.

### Ethical Considerations

The study was approved by the institutional review board of the National Institute of Preventive and Social Medicine (NIPSOM) (NIPSOM/IRB/2014/62; Dated: 24.08.2014). Informed assent was taken from the participants, and consents were taken from their parents before inclusion. All procedures were conducted following the guidelines laid out in the Declaration of Helsinki.

## Results

The study included 1,056 adolescents (mean age 14.95±0.78 years and 66.76% male) equally distributed between low and high-salinity areas (Table 1). Marked disparities in parental education were observed: in low salinity areas, 71.02% of mothers and 74.62% of fathers had secondary education or higher, compared to only 27.46% and 30.68% respectively in high salinity areas. Drinking water sources differed substantially by location, with deep tubewell predominating in low salinity areas (86.17%) versus pond water in high salinity areas (60.98%).

**Table 1.** Participant characteristics by residence.

| Characteristic | Overall<br>N = 1,056<sup>1</sup> | Residence |  |
| --- | --- | --- | --- |
|  |  | Low Salinity Area<br>N = 528<sup>1</sup> | High Salinity Area<br>N = 528<sup>1</sup> |
| **Age (years)** | 14.95 (0.78) | 14.95 (0.75) | 14.95 (0.81) |
| **Sex** |  |  |  |
| Male | 705 (66.76) | 352 (66.67) | 353 (66.86) |
| Female | 351 (33.24) | 176 (33.33) | 175 (33.14) |
| **Extracurricular Activity** |  |  |  |
| Don't do anything | 695 (65.81) | 370 (70.08) | 325 (61.55) |
| Pity works in a family | 321 (30.40) | 143 (27.08) | 178 (33.71) |
| Works in cultivator land | 33 (3.13) | 15 (2.84) | 18 (3.41) |
| Business | 7 (0.66) | 0 (0.00) | 7 (1.33) |
| **Mother's education** |  |  |  |
| Secondary | 520 (49.24) | 375 (71.02) | 145 (27.46) |
|  |  | Low Salinity Area<br>N = 528<sup>1</sup> | High Salinity Area<br>N = 528<sup>1</sup> |
| Primary | 242 (22.92) | 74 (14.02) | 168 (31.82) |
| Higher secondary or<br>Higher | 152 (14.39) | 68 (12.88) | 84 (15.91) |
| No formal education | 142 (13.45) | 11 (2.08) | 131 (24.81) |
| **Mother's occupation** |  |  |  |
| Household work | 980 (92.80) | 506 (95.83) | 474 (89.77) |
| Part-time employment | 50 (4.73) | 20 (3.79) | 30 (5.68) |
| Full-time employment | 21 (1.99) | 1 (0.19) | 20 (3.79) |
| Business | 5 (0.47) | 1 (0.19) | 4 (0.76) |
| **Father's education** |  |  |  |
| Higher secondary or<br>Higher | 556 (52.65) | 394 (74.62) | 162 (30.68) |
| Secondary | 370 (35.04) | 132 (25.00) | 238 (45.08) |
| No formal education | 78 (7.39) | 2 (0.38) | 76 (14.39) |
| Primary | 52 (4.92) | 0 (0.00) | 52 (9.85) |
| **Father's occupation** |  |  |  |
| Business/Skilled labor<br>/Shopkeeper | 755 (71.50) | 409 (77.46) | 346 (65.53) |
| Service/Teacher | 222 (21.02) | 113 (21.40) | 109 (20.64) |
| Farmer | 69 (6.53) | 5 (0.95) | 64 (12.12) |
| Daily labor/Rickshaw<br>puller | 10 (0.95) | 1 (0.19) | 9 (1.70) |
| **Monthly Family Income**<br>**(BDT)** | 8,400.00 (8,000.00,<br>10,000.00) | 8,500.00 (8,000.00,<br>9,500.00) | 8,000.00 (7,000.00,<br>11,000.00) |
| **Source of drinking**<br>**water** |  |  |  |
| Deep tubewell | 567 (53.69) | 455 (86.17) | 112 (21.21) |
| Pond | 322 (30.49) | 0 (0.00) | 322 (60.98) |
| Rainwater | 94 (8.90) | 0 (0.00) | 94 (17.80) |

| Characteristic | Residence |  |  |
| --- | --- | --- | --- |
|  | Overall<br>N = 1,056<sup>1</sup> | Low Salinity Area<br>N = 528<sup>1</sup> | High Salinity Area<br>N = 528<sup>1</sup> |
| Shallow tubewell | 54 (5.11) | 54 (10.23) | 0 (0.00) |
| Ring well/ Dug well | 19 (1.80) | 19 (3.60) | 0 (0.00) |
| **Source of cooking water** |  |  |  |
| Deep tubewell | 557 (52.75) | 455 (86.17) | 102 (19.32) |
| Pond | 399 (37.78) | 0 (0.00) | 399 (75.57) |
| Shallow tubewell | 54 (5.11) | 54 (10.23) | 0 (0.00) |
| Rainwater | 27 (2.56) | 0 (0.00) | 27 (5.11) |
| Ring well/ Dug well | 19 (1.80) | 19 (3.60) | 0 (0.00) |

<sup>1</sup>Mean (SD); n (%); Median (Q1, Q3)

Mean BMI was significantly higher in low salinity areas (22.56±1.46 kg/m^2^) compared to high salinity areas (17.58±2.14 kg/m^2^, p<0.001). Nutritional status differed markedly with 48.86% of participants in low salinity areas being overweight versus less than 1% in high salinity areas, while underweight prevalence was substantially higher in high salinity areas (23.67% vs. 0.19%). Family history of hypertension (97.73% vs. 82.77%, p<0.001) and extra salt intake habit (99.62% vs. 83.14%, p<0.001) were significantly more prevalent in low salinity areas (Table 2).

**Table 2:**
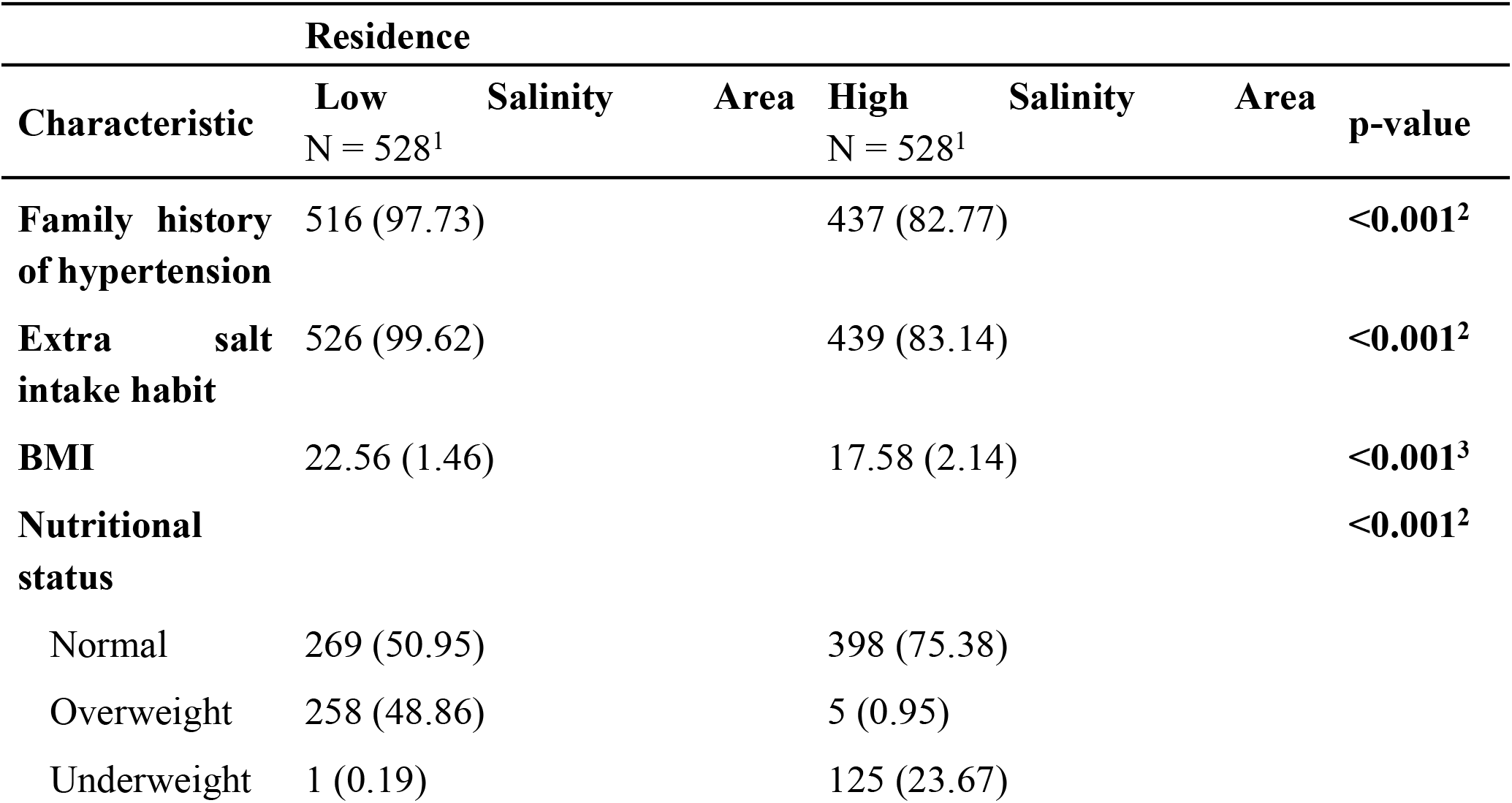
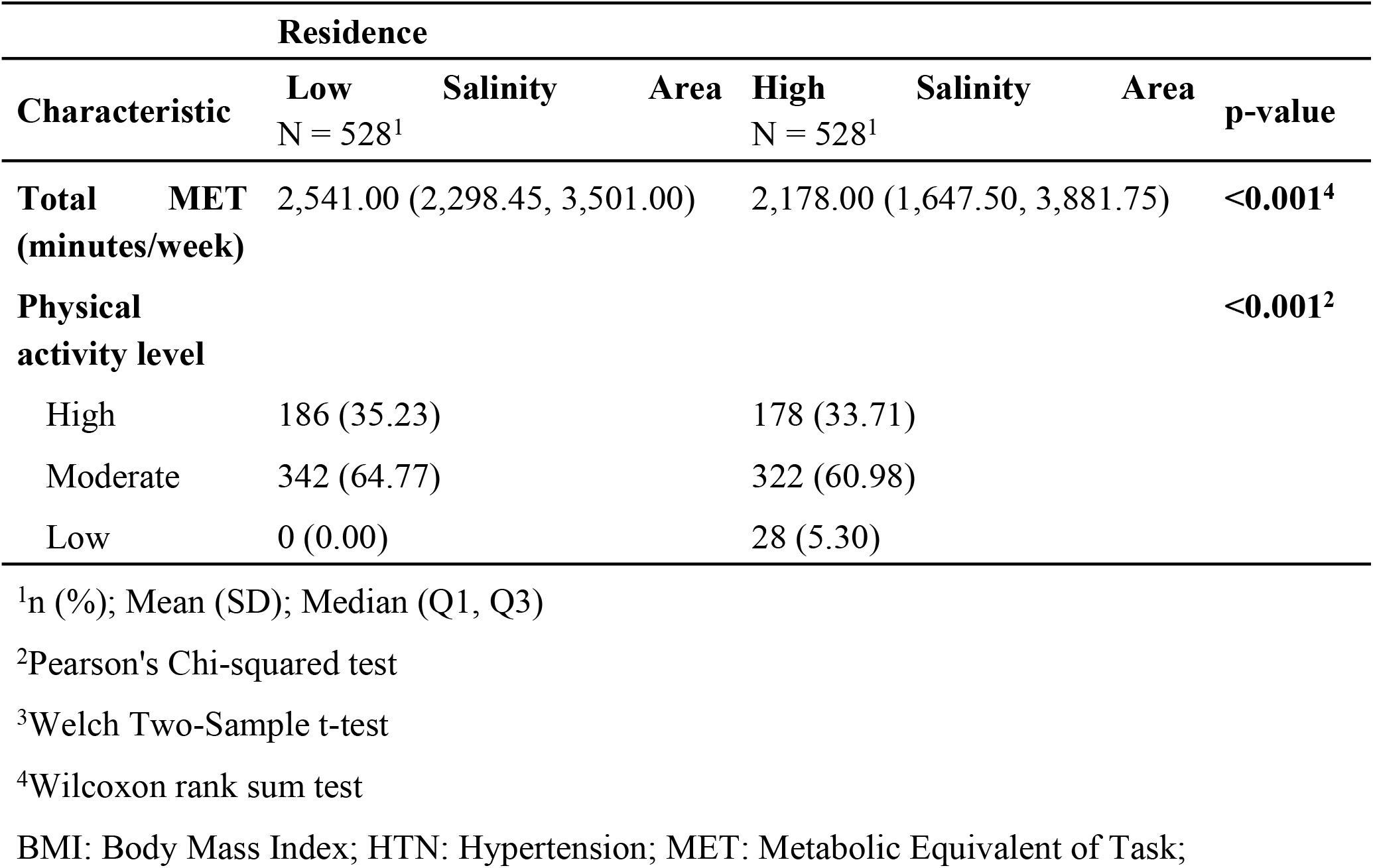
Distribution of Known Risk Factors of Hypertension by Residence.

Males consistently exhibited significantly higher blood pressure than females in both locations (all p<0.001). BMI showed strong positive correlations with both SBP and DBP in both areas (all p<0.001), with correlation coefficients ranging from 0.198 to 0.341. Age correlated with SBP only in low salinity areas (r=0.147, p<0.001) but not in high salinity areas (p=0.946). Log-transformed MET minutes also showed correlation with SBP in high-salinity areas (r=0.194, p<0.001) and with DBP in low-salinity areas (r=0.093, p=0.032) (Table 3).

**Table 3:**
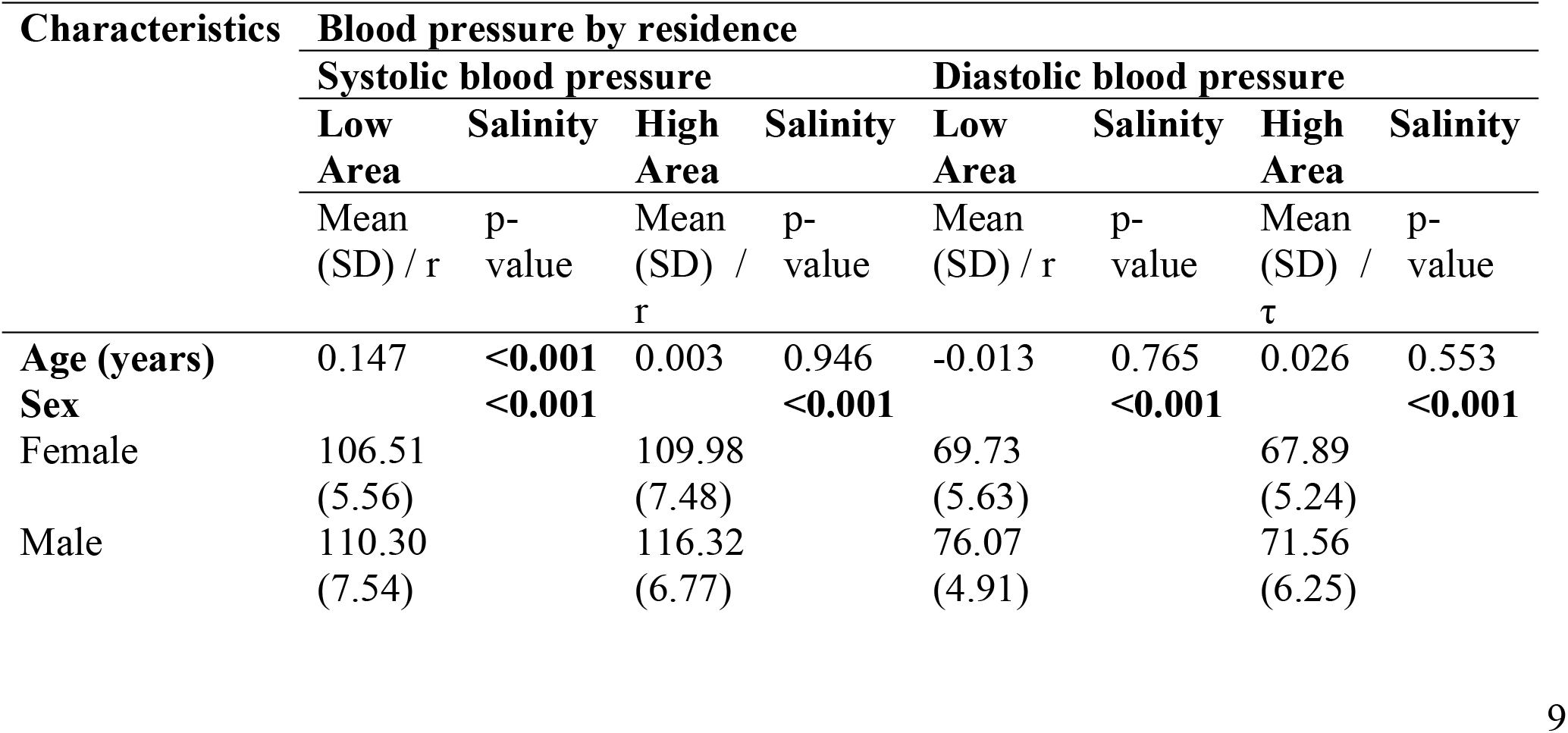
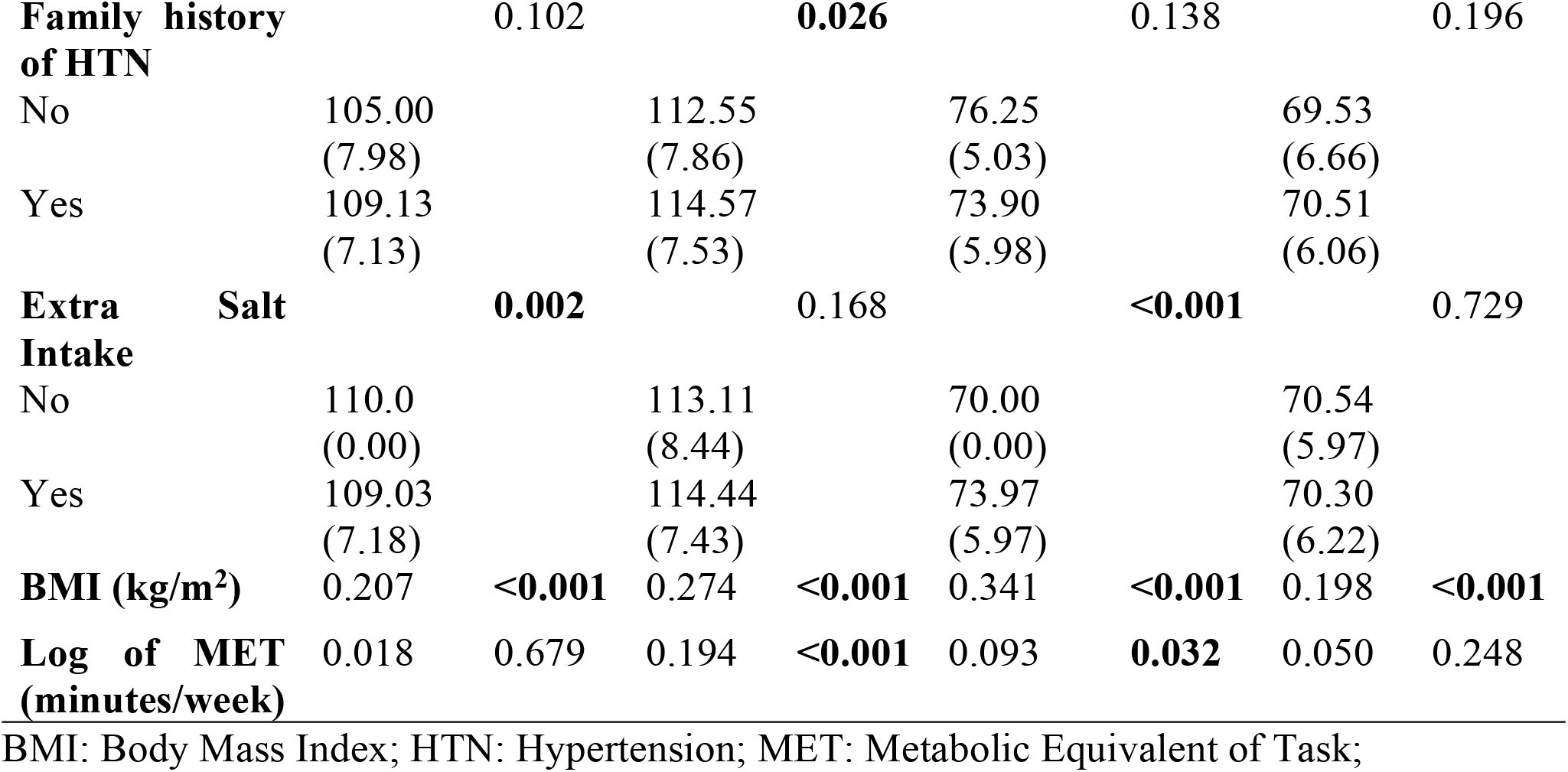
Association of Blood Pressure with Participant Characteristics.

| Characteristics | Blood pressure by residence |  |  |  |  |  |  |  |
| --- | --- | --- | --- | --- | --- | --- | --- | --- |
|  | Systolic blood pressure |  |  |  | Diastolic blood pressure |  |  |  |
|  | Low | Salinity | High | Salinity | Low | Salinity | High | Salinity |
|  | Area |  | Area |  | Area |  | Area |  |
|  | Mean | p- | Mean | p- | Mean | p- | Mean | p- |
|  | (SD) / r | value | (SD) / | value | (SD) / r | value | (SD) / | value |
| | | | r | | | | $\tau$ | |
| **Age (years)** | 0.147 | **<0.001** | 0.003 | 0.946 | -0.013 | 0.765 | 0.026 | 0.553 |
| **Sex** |  | **<0.001** |  | **<0.001** |  | **<0.001** |  | **<0.001** |
| Female | 106.51 |  | 109.98 |  | 69.73 |  | 67.89 |  |
|  | (5.56) |  | (7.48) |  | (5.63) |  | (5.24) |  |
| Male | 110.30 |  | 116.32 |  | 76.07 |  | 71.56 |  |
|  | (7.54) |  | (6.77) |  | (4.91) |  | (6.25) |  |
| **Family history of HTN** |  | 0.102 |  | **0.026** |  | 0.138 |  | 0.196 |
| No | 105.00<br>(7.98) |  | 112.55<br>(7.86) |  | 76.25<br>(5.03) |  | 69.53<br>(6.66) |  |
| Yes | 109.13<br>(7.13) |  | 114.57<br>(7.53) |  | 73.90<br>(5.98) |  | 70.51<br>(6.06) |  |
| **Extra Salt Intake** |  | **0.002** |  | 0.168 |  | **<0.001** |  | 0.729 |
| No | 110.0<br>(0.00) |  | 113.11<br>(8.44) |  | 70.00<br>(0.00) |  | 70.54<br>(5.97) |  |
| Yes | 109.03<br>(7.18) |  | 114.44<br>(7.43) |  | 73.97<br>(5.97) |  | 70.30<br>(6.22) |  |
| **BMI (kg/m<sup>2</sup>)** | 0.207 | **<0.001** | 0.274 | **<0.001** | 0.341 | **<0.001** | 0.198 | **<0.001** |
| **Log of MET (minutes/week)** | 0.018 | 0.679 | 0.194 | **<0.001** | 0.093 | **0.032** | 0.050 | 0.248 |

BMI: Body Mass Index; HTN: Hypertension; MET: Metabolic Equivalent of Task;

Both SBP and DBP differed significantly between locations (both p<0.001). High salinity areas showed higher median SBP (114.2±7.6 vs. 109.0±7.2 mmHg) but lower median DBP (70.3±6.2 vs. 74.0±6.0 mmHg) compared to low salinity areas, representing differences of approximately 5.2 mmHg and 3.7 mmHg, respectively (Figure 1).

**Figure 1:**
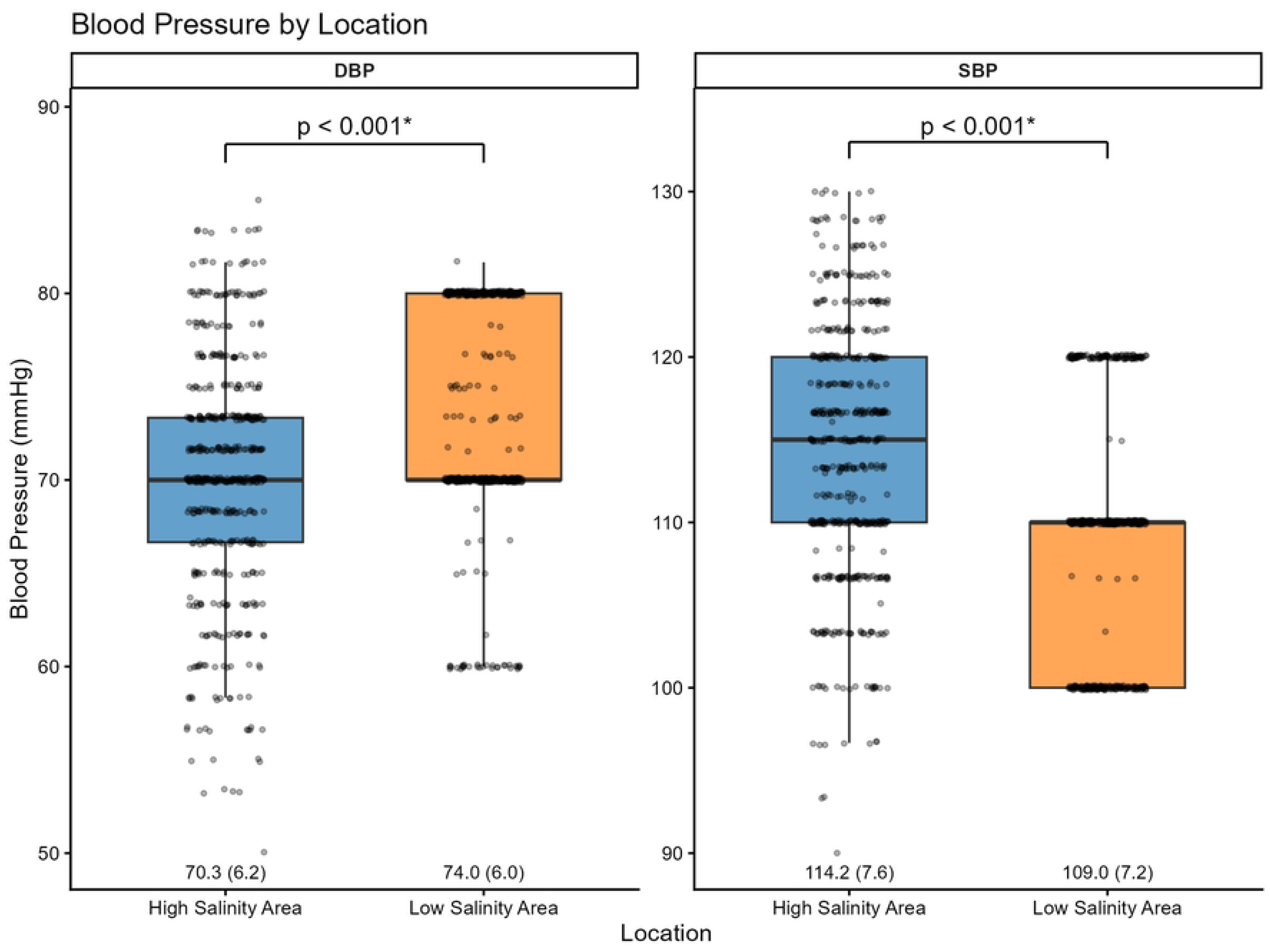
Blood Pressure by Residence.

After adjusting for age, sex, family history of hypertension, extra salt intake, BMI, and physical activity, residence in high salinity areas was independently associated with an 8.74 mmHg increase in SBP (95% CI: 7.22 to 10.26, p<0.001) compared to low salinity areas. No significant association was observed with DBP (β=-0.97 mmHg, 95% CI: -2.18 to 0.25, p=0.120) (Table 4). See S1 Table for the full model with coefficients (S1 File).

**Table 4:**
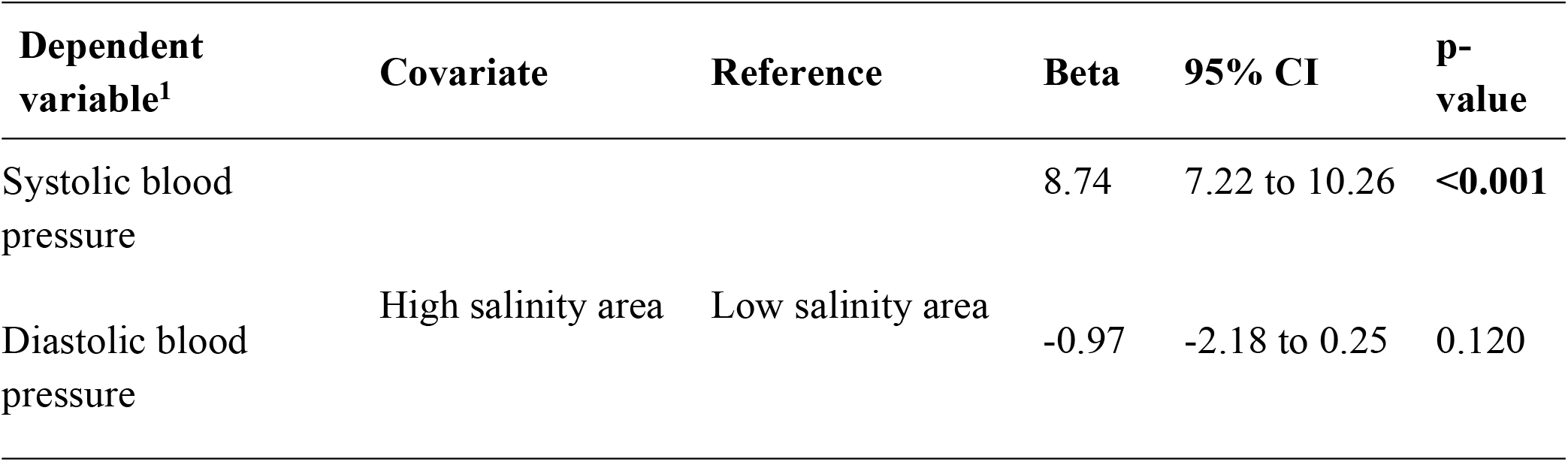
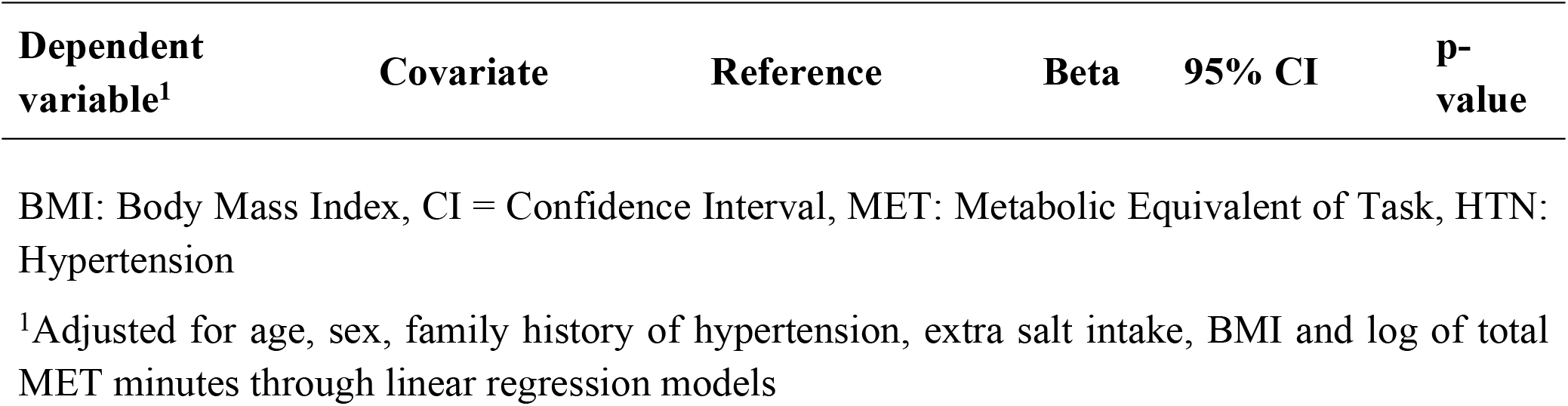
Effect of Life-Long Residence on Blood Pressure of Adolescents after Adjustment of Other Factors.

Sensitivity analysis using median quintile regression also showed a significant association of residence with salinity, with a high-salinity area being associated with higher median systolic blood pressure and lower diastolic blood pressure (S2 Table) (S1 File).

## Discussion

This cross-sectional comparative study examined blood pressure levels among school-going adolescents residing in high- and low-salinity areas of Bangladesh since birth. The findings reveal that after adjusting for established risk factors, life-long residence in high-salinity areas was independently associated with significantly higher systolic blood pressure among adolescents, although diastolic blood pressure showed no significant association. These results add to the growing body of evidence linking environmental salinity exposure to cardiovascular health outcomes in vulnerable populations.

The most striking finding from this study is the 8.74 mmHg increase in systolic blood pressure associated with residence in high-salinity areas after controlling for age, sex, family history of hypertension, extra salt intake, BMI, and physical activity. This magnitude of difference is clinically significant, as even modest increases in blood pressure during adolescence can have lasting implications. Blood pressure levels established in childhood and adolescence tend to track into adulthood, making early identification and intervention crucial for preventing future cardiovascular disease [16]. Recent meta-analytic evidence from 27 observational studies involving over 74,000 participants has demonstrated that higher drinking water salinity is associated with elevated systolic and diastolic blood pressure, particularly among coastal populations[23]. Our findings align with this broader pattern, though the magnitude of effect we observed for systolic blood pressure is notably larger than the pooled mean differences of 3.22 mmHg reported in the recent systematic review, possibly reflecting the particularly high salinity exposure in our study area and the vulnerability of our adolescent population.

The observed paradox of elevated systolic blood pressure but lower diastolic blood pressure in high-salinity areas compared to low-salinity areas warrants careful interpretation. While systolic blood pressure showed a clear positive association with salinity exposure, the crude comparison revealed lower diastolic blood pressure in high-salinity areas. This seemingly contradictory finding can be explained by the marked differences in nutritional status between the two populations. Participants from low-salinity areas had significantly higher BMI and a substantially greater prevalence of overweight status, which are well-established risk factors for elevated blood pressure [14]. The strong positive correlations between BMI and both systolic and diastolic blood pressure observed in our study support this interpretation. After adjusting for BMI and other confounders in the regression analysis, the association between salinity exposure and diastolic blood pressure became non-significant, suggesting that the crude differences in diastolic blood pressure were largely driven by differences in body composition rather than salinity exposure per se.

The mechanism by which environmental salinity exposure affects blood pressure is primarily mediated through increased sodium intake via drinking water. Coastal Bangladesh experiences particularly severe salinity intrusion, with sodium concentrations in groundwater reaching levels of 2,000 mg/L in some areas, far exceeding the WHO recommended guideline value of 200 mg/L for drinking water [12]. The high-salinity areas in our study primarily relied on pond water as their drinking water source, which is particularly vulnerable to saltwater contamination through tidal surges and soil runoff [6]. Chronic exposure to high-sodium drinking water leads to sodium retention, increased plasma volume, and arterial constriction, ultimately resulting in elevated blood pressure [17]. Additionally, excessive sodium intake can reduce endothelial nitric oxide production, increase vascular stiffness, and modulate autonomic nervous system activity, all of which contribute to hypertension development [23].

Our findings are consistent with previous research conducted in coastal Bangladesh. Talukder and colleagues found that young adults in coastal Bangladesh consuming highly saline drinking water above 600 mg/L had significantly higher systolic and diastolic blood pressure compared to those consuming water with lower salinity levels [13]. Similarly, Scheelbeek and colleagues demonstrated in a cohort study that drinking water salinity was associated with raised blood pressure in coastal populations [9]. The consistency of these findings across different age groups and study designs strengthens the evidence for a causal relationship between environmental salinity exposure and elevated blood pressure. Interestingly, some research has shown that the relationship between drinking water salinity and blood pressure may be modified by other minerals present in the water. A study by Naser and colleagues found that mild-salinity water containing higher concentrations of calcium and magnesium was associated with lower blood pressure, suggesting that these minerals may have protective effects [24]. However, this potential protective effect appears to be limited to specific mineral compositions and does not negate the overall harmful effects of high sodium exposure.

The observed differences in participant characteristics between high- and low-salinity areas highlight important contextual factors that influence both salinity exposure and health outcomes. Nearly one-fourth of the participants from high salinity areas were found to be underweight, consistent with the findings of Mazumder and colleagues [25]. Nearly half of the participants were found to be overweight in the low-salinity areas. The marked nutritional disparities between the two areas, with high rates of underweight in high-salinity areas versus high rates of overweight in low-salinity areas, reflect broader socioeconomic differences that may independently affect cardiovascular health. These findings underscore that salinity exposure does not occur in isolation but is part of a complex web of environmental, social, and economic factors that shape population health in coastal Bangladesh.

The universal prevalence of extra salt intake in low-salinity areas compared to high-salinity areas is another noteworthy finding. This likely reflects dietary patterns and food preservation practices that vary by region and socioeconomic status. The high prevalence of extra salt intake in low-salinity areas may partially offset the protective effect of lower environmental salinity exposure, though our regression analysis attempted to account for this factor. The higher prevalence of family history of hypertension in low-salinity areas could reflect better diagnosis and awareness of hypertension in areas with greater healthcare access, or it could indicate a genuine genetic predisposition that contributes to the overall blood pressure patterns observed.

The positive correlation between physical activity and systolic blood pressure observed in high-salinity areas, while statistically significant, is counterintuitive given that physical activity is generally associated with lower blood pressure. This finding may reflect residual confounding from obesity or measurement related to how physical activity was assessed in this population.

This study has several important implications for public health policy and practice in Bangladesh and other low-lying coastal regions vulnerable to climate change. First, our findings highlight the urgent need for interventions to provide safe drinking water to coastal populations. Climate change projections suggest that saltwater intrusion will intensify in coming decades, potentially affecting millions more people globally [6]. Nature-based solutions such as rainwater harvesting, pond sand filters, and managed aquifer recharge systems offer potential strategies to reduce drinking water salinity, though these require further evaluation of their effectiveness and implementation feasibility in real-world settings [23]. Second, screening programs for hypertension should be prioritized in coastal areas with high environmental salinity exposure, particularly among children and adolescents who may benefit most from early intervention. Third, health education campaigns should raise awareness about the cardiovascular risks associated with saline water consumption and promote behavioral strategies to minimize sodium intake from all sources.

The study had several limitations that need to be acknowledged. The cross-sectional design precludes definitive causal inference, though the biological plausibility of the association and consistency with longitudinal studies strengthen confidence in our findings. We relied on single-point-in-time blood pressure measurements, which may not fully capture individuals’ typical blood pressure levels, though we used standard protocols, including multiple readings, to minimize measurement error. Information on total sodium intake was not directly measured but rather inferred from water sources and self-reported dietary practices, which may not accurately reflect actual sodium consumption. We did not measure drinking water salinity levels directly in this study but rather relied on previous surveys to classify areas as high- or low-salinity regions, which may not account for temporal or spatial variation in salinity within these areas. The marked socioeconomic differences between study areas introduce the possibility of residual confounding despite our statistical adjustments. Finally, the study was conducted in specific villages in Bangladesh, which may limit generalizability to other coastal populations with different environmental conditions, dietary patterns, and genetic backgrounds. Despite these limitations, this study makes important contributions to understanding the health impacts of environmental salinity exposure in adolescent populations. The use of a comparative design with careful selection of high- and low-salinity areas based on national survey data, comprehensive assessment of known hypertension risk factors, and rigorous statistical adjustment for confounders strengthens the validity of our findings. The focus on adolescents is particularly valuable given the limited research on this age group and the importance of early-life blood pressure patterns for long-term cardiovascular health.

Future research should employ longitudinal designs to establish temporal relationships between salinity exposure and blood pressure development over time. Studies should include direct measurement of drinking water salinity, total dietary sodium intake, and other relevant minerals such as calcium and magnesium that may modify the salinity-blood pressure relationship. Investigation of potential interventions to reduce salinity exposure or mitigate its health effects is urgently needed, particularly scalable and sustainable solutions appropriate for resource-limited coastal settings. Research should also explore whether genetic factors modify individual susceptibility to the blood pressure effects of environmental salinity exposure, which could inform targeted prevention strategies.

## Conclusion

In conclusion, this study provides compelling evidence that life-long residence in high-salinity areas of coastal Bangladesh is independently associated with elevated systolic blood pressure among adolescents, after accounting for multiple established risk factors. These findings highlight environmental salinity exposure as an important and modifiable risk factor for hypertension in coastal populations. Given the projected intensification of saltwater intrusion due to climate change and the millions of people worldwide who rely on saline-affected water sources, addressing this environmental health threat should be a priority for global health and climate adaptation efforts. Integrated approaches that combine provision of safe drinking water, hypertension screening and management, health education, and sustainable environmental management will be essential to protect the cardiovascular health of current and future generations in coastal regions.

## Acknowledgement

We acknowledge the school authorities and parents for their support during the data collection. We are grateful to the adolescents who voluntarily participated in the study.

## Use of generative AI

The Large Language Model Claude AI 4.5 Sonnet was used during the preparation of the manuscript to ensure correct grammar and flow of writing, as the authors are not native speakers of English. Any output from the AI model was thoroughly reviewed, and the authors take full responsibility for the final results.

## Data availability

An anonymized dataset is available as a supplementary file (S1 Data).

## Additional Information

**S1 Fig. Post-hoc analysis of the linear regression model for systolic blood pressure**

**S2 Fig. Post-hoc analysis of the linear regression model for diastolic blood pressure**

**S1 File. Supplementary tables**

**S1 STROBE Checklist. STrengthening the Reporting of OBservational studies in Epidemiology (STROBE) checklist for cross-sectional studies**

